# Ventilation heterogeneity is increased in adults exposed to coal mine fire-related PM_2.5_

**DOI:** 10.1101/2023.06.08.23291105

**Authors:** Thomas McCrabb, Brigitte Borg, Caroline X. Gao, Catherine Smith, David Brown, Jillian Ikin, Annie Makar, Tyler Lane, Michael J. Abramson, Bruce R Thompson

## Abstract

**Background and Objectives:** The Hazelwood Health Study was set up to study long term health effects of a mine fire that blanketed residents of the Latrobe Valley with smoke for 45 days in 2014. The Respiratory Stream specifically assessed the impact of fine particulate matter <2.5μm diameter (PM_2.5_) exposure from mine fire smoke on lung health. The multiple breath nitrogen washout (MBW) test assesses ventilation heterogeneity, which may detect early airways dysfunction not identified using standard tests such as spirometry. This analysis assessed the association of PM_2.5_ exposure with measures of ventilation heterogeneity.

**Methods:** Exposed (Morwell) and unexposed (Sale) participants were recruited 3.5-4 years after the fire from those who had participated in an Adult Survey. MBW was performed to measure lung clearance index (LCI), functional residual capacity (FRC), acinar (Sacin) and conductive (Scond) ventilation heterogeneity. PM_2.5_ exposure was estimated with emission and chemical transport models. Multivariate linear regression models were fitted controlling for confounders.

**Results:** We recruited 519 participants. MBW tests were conducted on 504 participants with 479 acceptable test results (40% male; 313 exposed, 166 unexposed). Exposure to mine fire-related PM_2.5_ was associated with increasing Scond (β=2.15/kL, 95%CI: 0.67-3.63, p=0.006), which was comparable to the estimated effect on Scond of 7.9 years of aging. No other MBW outcomes were significant.

**Conclusions:** Increasing exposure to PM_2.5_ was associated with increased ventilation heterogeneity in the conductive region of the lungs 4 years after the event.

## Introduction

In 2014, a fire in the Hazelwood coal mine, in south-eastern Australia, blanketed the nearby town of Morwell in smoke for 45 days. The National Environment Protection Measure for fine particles < 2.5μm diameter (PM_2.5_) was exceeded on 27 days.^1^ In response to community concerns around possible health impacts of the smoke on local residents, the Victorian State Government established the Hazelwood Health Study (HHS) to formally investigate long term health effects.

The Respiratory Stream of the HHS was set up to identify whether people exposed to the smoke from the mine fire have or develop over time clinical or sub-clinical respiratory conditions that may be associated with clinically important adverse health consequences in the future, compared to those who had minimal or no exposure.

In 2017-18, 3.5 to 4 years after the mine fire, the Respiratory Stream collected clinical data in Morwell (exposed) and Sale (unexposed). Findings reported so far included: asthmatics in Morwell had worse asthma control than those from Sale.^2^ There was an association between PM_2.5_ exposure and chest tightness and chronic cough particularly in smokers^3^ and spirometric features of COPD in non-smokers.^3^ There was also an association between PM_2.5_ exposure and worsening lung mechanics measured with oscillometry.^4^

Presentation and diagnosis of respiratory symptoms can take years following exposure to PM_2.5_.^5^ This may in part be due to the measurement tools used to monitor changes in physiology. Spirometry, the most common technique for assessing ventilatory function, is not particularly sensitive to detecting early changes in peripheral airway function, and disease may be quite advanced before abnormalities are detected. Multiple breath washout (MBW) is potentially a more sensitive and discriminating method in assessing peripheral airways dysfunction.^5^ In a normal healthy lung, gas mixing should be even across the lung. In early lung pathology predominantly affecting peripheral airways, gas mixing becomes less efficient.^6^ MBW assesses gas mixing (known as ventilation heterogeneity) within the lungs and hence may be a more appropriate tool for detecting early changes in airway function.

This cross-sectional analysis aimed to detect an association between mine fire-related PM_2.5_ exposure and changes in the ventilation heterogeneity. If present, an association could be an early marker of small airways disease in the exposed population.

## Methods

### Study design and setting

This cross-sectional analysis studied data collected in the longitudinal Hazelwood Adult Study Respiratory stream.

### Participants and recruitment

Participants in this study were a sub-sample of the HHS Adult Survey participants (conducted 2016 and 2017).^7^ A total of 3096 Morwell and 960 Sale residents provided responses in HHS Adult Survey. Sale was selected as a control group due to its similar rural setting, age distributions and socio-economic factors, and minimal exposure to the coal mine fire smoke.^8^ Respondents were randomly selected to participate in the respiratory stream with those reporting current asthma medication or attacks of asthma oversampled. Detailed procedures have been published elsewhere.^2,4^ In summary, clinical testing occurred in Morwell from August to December 2017 and Sale from January to March 2018. Pulmonary function tests (PFT), interviewer administered questionnaires and anthropometric measurements were collected by trained respiratory scientists. Study data were collected and managed using REDCap electronic data capture tool hosted and managed by Helix (Monash University)^9^.

Spirometry and MBW (nitrogen washout method) were performed on the EasyOne Pro™ Lab (ndd Medizintechnik AG, Zürich, CH) following the American Thoracic Society (ATS) and European Respiratory Society (ERS) guidelines^10^ and the consensus statement for inert gas washout measurements.^11^

Standardization between staff and testing equipment was achieved with detailed study protocols and quality control procedures. This study obtained ethics approval from the Monash University Human Research Ethics Committee (project number: 1078) and the Alfred Ethics Committee (project number: 90/21). All participants provided written informed consent.

### Measures

#### Exposure – PM_2.5_

Due to a lack of ground-level air pollution monitoring at the early stage of the mine fire, retrospective modelling of the mine fire-related PM_2.5_ concentration was conducted by the Commonwealth Scientific and Industrial Research Organisation (CSIRO) Oceans & Atmosphere.^8^ The modelled spatial and temporal distribution of mine fire-related PM_2.5_ were linked to time-location diary data collected as a part of the Adult Survey to estimate individual-level mean daily mine fire-related PM_2.5_ exposure over the fire period.^12^

#### MBW Outcomes

MBW measures the efficiency of gas mixing in the lungs with four outcomes: Lung Clearance Index (LCI), Functional Residual Capacity (FRC), Sacin and Scond. Lung Clearance Index (LCI) is a global measure of ventilation heterogeneity, which measures the number of lung turnovers (TO) to eliminate nitrogen from the lungs. Functional Residual Capacity (FRC) is the end expiratory lung volume during tidal breathing. The analysis of the slope of the normalised nitrogen alveolar plateau (phase III slope) was performed to distinguish ventilation heterogeneity associated with the acinar (respiratory) zone (measured by Sacin) and the conductive zone (measured by Scond) of the lung. LCI and FRC were reported as the average of acceptable tests. Scond and Sacin were also reported as the average of acceptable tests and calculated^13^ using WBreath research analysis software (ndd Medizintechnik AG, Zürich, Switzerland).

#### Covariates

In this study we considered a range of potential confounders. Self-reported sex, employment history, highest level of completed education and occupational exposures (employment in dusty or polluted environments for at least 6 months) collected in the Adult Survey.^7^ Age, ethnicity, height and weight were recorded during clinical testing. Smoking status and self-reported asthma were collected via interviewer administered surveys based on modified versions of the European Community Respiratory Health Survey (ECRHS).^14^ Spirometric Chronic Obstructive Pulmonary Disease (COPD) was defined as post-bronchodilator FEV_1_/FVC z score < -1.64.^3,15,16^

### Statistical Analysis

Descriptive statistics were obtained for participant characteristics and clinical outcomes for four exposure groups: (1) non-exposed (Sale) participants, (2) low, (3) medium, and (4) high mine-fire smoke exposure. The low, medium and high exposure groups were assigned according to the tertiles of mine fire-related PM_2.5_ concentrations in Morwell. Crude statistical comparisons between groups were made using Pearson χ^2^ tests for categorical measures and one-way analysis of variance for continuous measures.

Multivariate linear regression models were fitted to evaluate associations between PM_2.5_ exposure and individual outcomes, controlling for key confounders (see above). Statistical weights were developed to account for oversampling of asthmatics. To ensure the robustness of the findings, three models were fitted for each outcome: (1) unweighted complete cases model (missing data excluded), (2) unweighted and imputed model, and (3) weighted and imputed model. All models were repeated including or excluding the residential location (Morwell/Sale) as a potential confounding factor which was also related to exposure.

Statistical analyses were performed using Stata version 16 (Stata Corporation, College Station, Texas 2016).

## Results

Altogether 519 adults (346 from Morwell and 173 from Sale) participated in the respiratory steam. MBW tests were conducted in 504 participants with 479 acceptable LCI and FRC test results (313 from Morwell and 166 from Sale). An additional two records were excluded for Sacin and 16 records for Scond due to not meeting acceptability criteria. Tests were not conducted for 15 participants (12 due to equipment issues and 3 due to contraindications to MBW testing).

Among the participants with acceptable MBW results, the characteristics were comparable between exposure groups, except for higher proportions of older and female participants in the non-exposed group (lower weight was likely to be a consequence of more female participants, see Table 1). There were some differences between participants who had acceptable MBW results and those who were not tested or had no acceptable results, who were older and less likely to be employed (see Table S1).

**Table 1.**
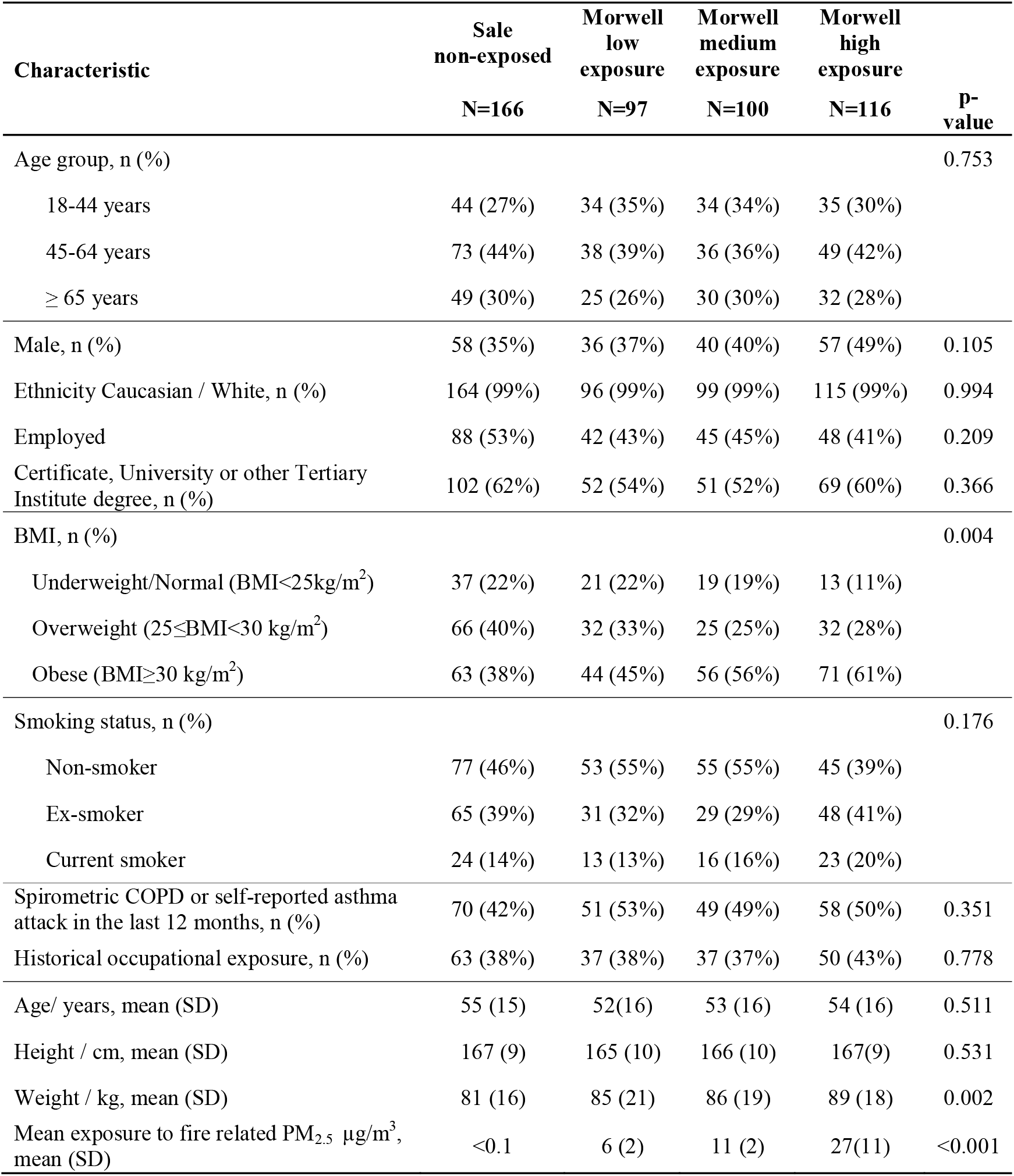
Characteristics of 479 participants who had acceptable MBW test results.

Distributions of LCI, FRC, Sacin and Scond are summarised by exposure groups in Figure 1 (descriptive statistics provided in Table S2). Participants from the unexposed group (Sale) had slightly higher unadjusted levels of FRC compared with exposed (Morwell) participants, which might be associated with heavier weight in Morwell participants. The unadjusted Sacin scores were also slightly higher in unexposed participants than low and medium exposed participants. On the other hand, unadjusted Scond was significantly higher in exposed than unexposed participants.

**Figure 1.**
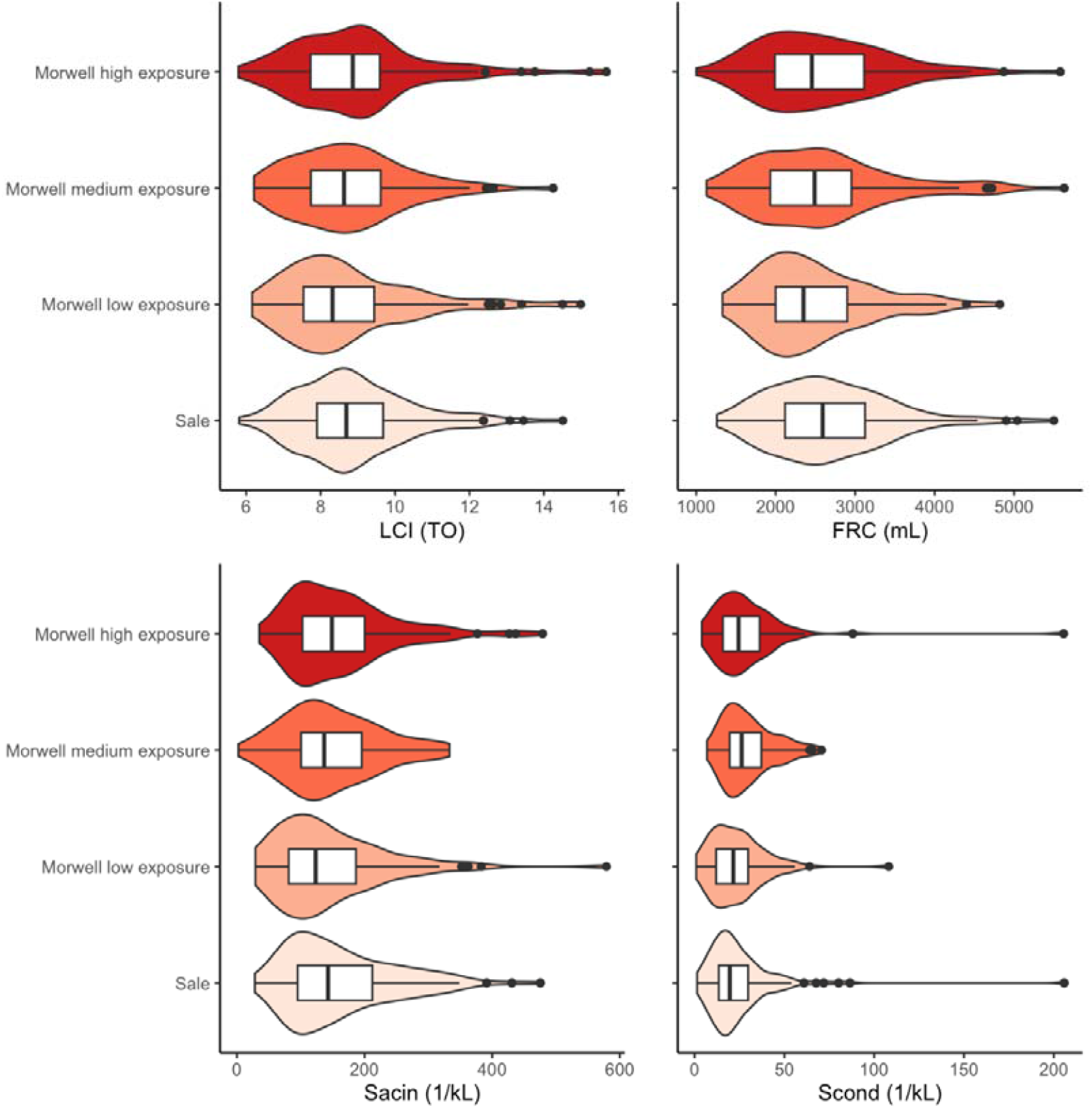
Distributions (boxplots and violin plots) of MBW outcomes by mine fire exposure groups. Note: Missing data include 40 records for LCI and FRC, 42 records for Sacin and 56 records for Scond. Abbreviations: LCI = Lung Clearance Index, FRC = Functional Residual Capacity, Sacin = acinar ventilation heterogeneity, Scond = conductive ventilation heterogeneity.

Multivariate linear regression models (Figure 2 and Table S3) suggested exposure to mine fire-related smoke was associated with higher Scond. For a 10 μg/m^3^ increase in mean mine fire-related PM_2.5_ exposure, there was a 2.15 per kL (95%CI 0.67, 3.63) increase in Scond, estimated from the unweighted and unimputed model without exposure site (Morwell/Sale) as a covariate, see Figure 2. This effect was comparable to about 7.9 years of ageing on SCond estimated from the same model (2.73 per kL difference in Scond associated with 10 years age difference), see Table S3. Consistent effects were estimated from imputed and/or weighted models with slightly smaller effects in weighted models, which primarily give lower weight to older participants who were more likely to participate in the respiratory stream testing.

**Figure 2.**
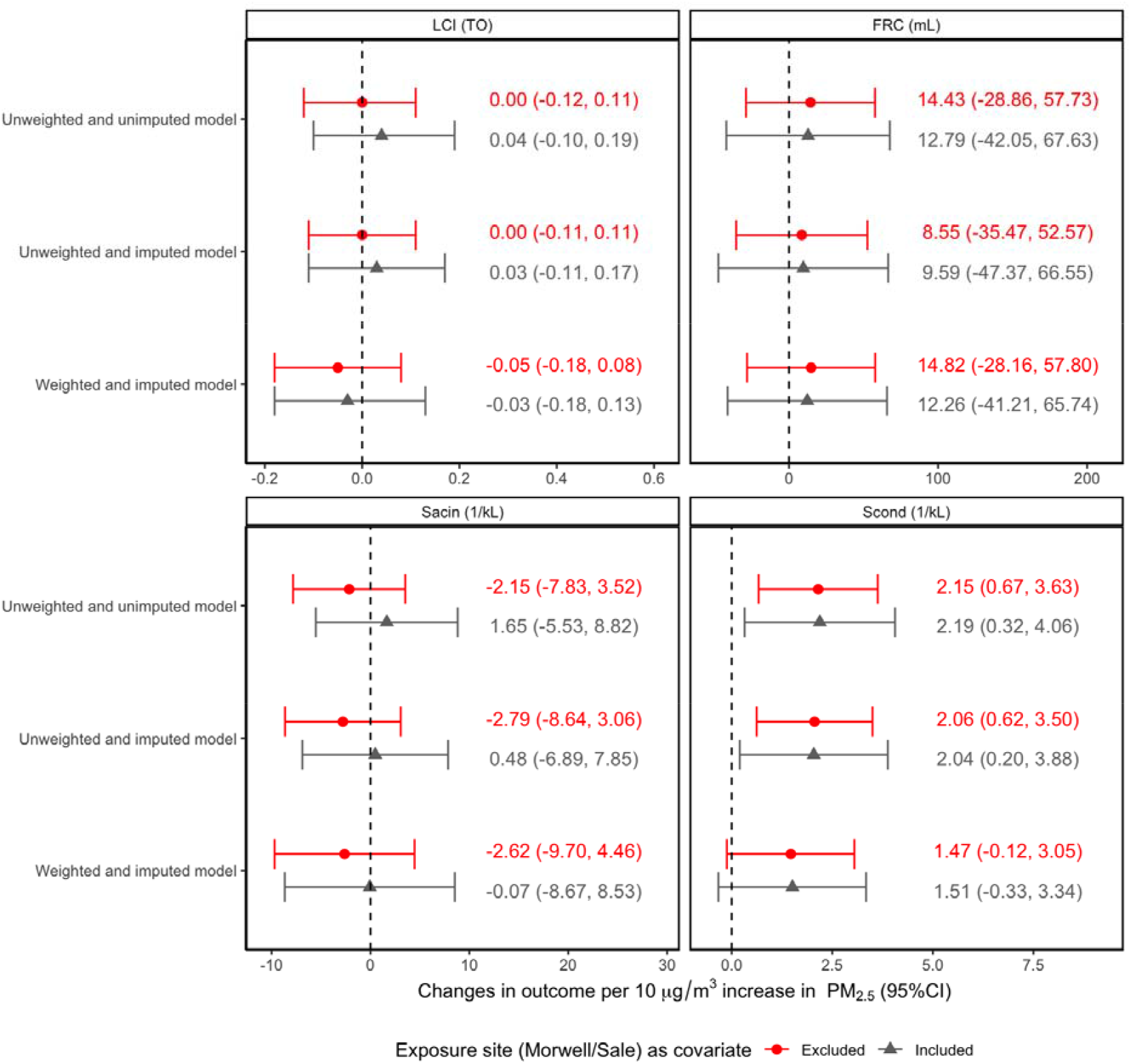
Outcomes associated with 10 μg/m^3^ increase in mean mine fire-related PM_2.5_ exposure estimated from different models. Note: all coefficients were estimated from multivariate linear regressions adjusting for age, sex, height, weight, employment, education, smoking, asthma or COPD and work exposure. Missing data included 40 records for LCI and FRC, 42 records for Sacin, 56 records for Scond and 6 records for education. Abbreviations: LCI = Lung Clearance Index, FRC = Functional Residual Capacity, Sacin = acinar ventilation heterogeneity, Scond = conductive ventilation heterogeneity

Although Sacin was not associated with PM_2.5_ exposure, it was higher in former and current smokers and those with spirometric COPD or a self-reported asthma attack in the last 12 months (Table S4).

## Discussion

A period of four years after the 2014 Hazelwood mine fire, we found PM_2.5_ exposure was associated with greater ventilatory heterogeneity in the small proximal conducting airways (Scond) of the lung, but no evidence of association with peripheral ventilatory heterogeneity (Sacin), FRC or LCI. The effect on Scond exceeded known age effects, which suggests exposure to coal mine fire-related PM_2.5_ led to premature lung ageing.^17^ To our knowledge, this is the first publication looking at the effects of PM_2.5_ exposure from a landscape fire (e.g., wildfire and mine fire) on ventilation heterogeneity using modern methods in adults.

The association of PM_2.5_ from an open cut coal mine fire and Scond increase is a novel finding. Negative outcomes related to PM_2.5_ exposure^18^ are motivating research into various PM_2.5_ sources such as wildfires, traffic pollution and residential indoor combustion. While wildfires may emit similar pollutants, the duration and intensity are different from the Hazelwood event – which may result in different physiological outcomes. MBW is an established clinical tool with the most commonly used outcome measure being the LCI.^6^ Current studies of ventilation heterogeneity partitioned by ventilatory region are aimed at early identification or interventions in patients with asthma,^19^ COPD,^20^ lung transplantation^21^ and cystic fibrosis^22^. Partitioning of ventilation heterogeneity by ventilatory region may provide further insights into lung pathophysiology due to PM_2.5_ exposure.

The changes seen in ventilatory heterogeneity in the current study agree with our previous analysis of airway mechanics in the same cohort. In that study,^4^ it was demonstrated that reactance in the peripheral airways (Xrs5) became more negative (worsened) with an increase in the corresponding area under the curve (AX5) as exposure to PM_2.5_ increased, suggesting airway closure commenced at a higher lung volume. Such changes in the airway closure may impact on Scond.^23^

The mechanism behind PM_2.5_ exposure increasing Scond is unclear, but there are a few possible explanations. In smokers with normal spirometry, increased bronchomotor (airway muscle) tone had a negative impact on Scond while not affecting Sacin.^24,25^ While the exposure here was shorter term and less intense than smoking, this provides a plausible mechanism for our findings. PM_2.5_ is small enough to deposit within the small airways proximal to the acinar entrance.^26^ The increase in Scond could mean particles are depositing within these airways, potentially affecting smooth muscle tone and convective gas mixing.^27^ Verbanck et al, showed normalisation of Scond in smokers with preserved spirometry on cessation of smoking over a few weeks to months,^28^ which might reflect recovery of airway inflammation. The association between increasing PM_2.5_ exposure and Scond may reflect increased bronchomotor tone or airway inflammation in the conductive region of the lung.

The lack of a significant change in Sacin suggests limited deposition in the peripheral airways. We cannot determine from this study why PM_2.5_ from the fire did not appear to have a detectable effect on Sacin. Based on evidence that Sacin was increased among smokers,^29,30^ we expected that PM_2.5_ exposure would increase Sacin heterogeneity because its small size would also allow it to reach the acinar region of the lungs. However, this was not supported by our findings. A possible reason for Sacin to not be altered in those exposed to the coal mine smoke may be due to overall particle size distribution. We know particles around 2.5 μm will deposit in airways proximal to the acinar entrance. Particles finer than 2 μm have been demonstrated to deposit peripheral to the acinar entrance by a process of diffusion. However, it has also been demonstrated that particles <1.8 μm have greater exhalation, ie. these particles are inhaled but then exhaled from the lung.^31^ It is possible that the overall particle size was not ultrafine and hence significant particle deposition in airways peripheral to the acinar entrance did not occur.

Although this study was not specifically designed to detect the effects of smoking on MBW outcomes, we confirmed prior evidence regarding impact of smoking on ventilation heterogeneity as we also found an association between increased Sacin and current and ex-smokers.^29,30^ These findings align with previous work on Sacin and suggests the measurements in the current study were valid. Sacin was influenced by tobacco smoke exposure, but not from the mine fire smoke, or at least not several years after the exposure.

### Strengths and limitations

A major strength was utilising an objective non-invasive clinical measurement that required minimal exertion from the participant following standardised procedures. A further strength was estimating individual PM_2.5_ exposure. Statistical models were fitted to adjust for known confounders with sample weighting and imputation of missing data to allow for comparison with complete case models.

However, this study also had some limitations. A lengthy (two-hour) clinical visit and reduced engagement from those with more symptoms might have biased the study. Relevant physiological (age, weight, height, sex) and demographic (occupation, education, smoking) cofounders were addressed. However, residual confounding may still have influenced the results. There were no data from before the mine fire and therefore we could not exclude pre-existing differences in ventilation heterogeneity. Although the effects of gaseous air pollutants could not be excluded, carbon monoxide was highly collinear with PM_2.5_.^8^ The findings might not be generalisable to culturally and linguistically more diverse communities exposed to landscape fire smoke elsewhere.

## Conclusion

There was a clear association between PM_2.5_ exposure from the coal mine fire smoke and increased heterogeneity in the conductive airways. This study adds to the limited literature on health effects of coal mine fire smoke exposure. More frequent similar events are expected as climate change accelerates. Future town planning should account for the health risks associated with decommissioned coal mines nearby. Further studies can build on these novel findings for early detection and treatment of those most affected. Follow-up investigations should identify the trajectory of ventilation heterogeneity and improve understanding of mechanisms from similar exposures.

## Supporting information

STROBE checklist

## Data Availability

The data are confidential and therefore not available to other researchers.

## Acknowledgments

The Hazelwood Health Study is funded by the Victorian Department of Health. However this paper presents the views of the authors and not those of the Department. We wish to thank the following people who assisted in conducting the study, but are not named authors: Susan Denny, Kylie Sawyer, Shantelle Allgood and Kristina Thomas. We wish to thank CSIRO for air pollution modelling and Danny Brazzale for reviewing the paper. Most of all, the study team would like to acknowledge the contributions of all community members who have participated in the study to date.

## Supplementary Material

**Table S1.** Differences between those who had acceptable MBW tests results and those who were not tested for MBW or had no acceptable results.

| Characteristic | With MBW<br>measure<br>N=479 | Without<br>MBW<br>measure<br>N=40 | p-value |
| --- | --- | --- | --- |
| Age group, n (%) |  |  | <0.001 |
| 18-44 years | 147 (31%) | 4 (10%) |  |
| 45-64 years | 196 (41%) | 14 (35%) |  |
| ≥ 65 years | 136 (28%) | 22 (55%) |  |
| Male, n (%) | 191 (40%) | 22 (55%) | 0.062 |
| Ethnicity Caucasian / White, n (%) | 474 (99%) | 40 (100%) | 0.516 |
| Employed | 223 (47%) | 6 (15%) | <0.001 |
| Certificate, University or other Tertiary Institute degree, n (%) | 274 (58%) | 17 (44%) | 0.085 |
| Body Mass Index (BMI), n (%) |  |  | 0.842 |
| Underweight/Normal (BMI<25 kg/m<sup>2</sup>) | 90 (19%) | 9 (23%) |  |
| Overweight (25≤BMI<30 kg/m<sup>2</sup>) | 155 (32%) | 12 (30%) |  |
| Obese (BMI≥30 kg/m<sup>2</sup>) | 234 (49%) | 19 (48%) |  |
| Smoking status, n (%) |  |  | 0.769 |
| Non-smoker | 230 (48%) | 19 (48%) |  |
| Ex-smoker | 173 (36%) | 13 (33%) |  |
| Current smoker | 76 (16%) | 8 (20%) |  |
| Spirometric COPD or self-reported asthma attack in the last 12 months, n (%) | 228 (48%) | 23 (57%) | 0.229 |
| Historical occupational exposure, n (%) | 187 (39%) | 18 (45%) | 0.459 |
| Age/ years, mean (SD) | 54(16) | 66 (15) | <0.001 |
| Height / cm, mean (SD) | 166 (9) | 165 (9) | 0.307 |
| Weight / kg, mean (SD) | 85 (18) | 82 (19) | 0.302 |
| Mean exposure to fire related PM<sub>2.5</sub> µg/m<sup>3</sup>, mean (SD) | 10 (12) | 13 (14) | 0.144 |

Missing data include 6 records for education level. SD = Standard Deviation

**Table S2.**
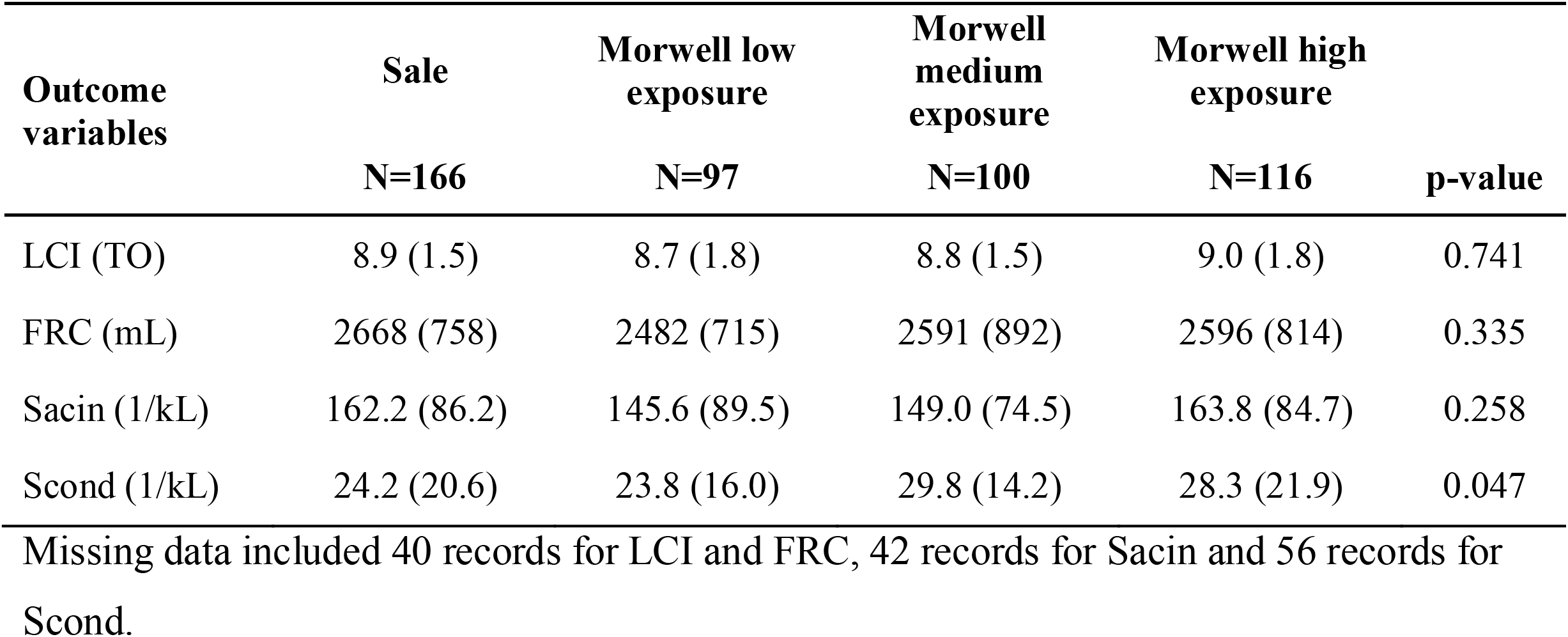
Distribution mean (SD) of outcomes by mine fire exposure groups.

| **Outcome variables** | **Sale** | **Morwell low exposure** | **Morwell medium exposure** | **Morwell high exposure** |  |
| --- | --- | --- | --- | --- | --- |
|  | **N=166** | **N=97** | **N=100** | **N=116** | **p-value** |
| LCI (TO) | 8.9 (1.5) | 8.7 (1.8) | 8.8 (1.5) | 9.0 (1.8) | 0.741 |
| FRC (mL) | 2668 (758) | 2482 (715) | 2591 (892) | 2596 (814) | 0.335 |
| Sacin (1/kL) | 162.2 (86.2) | 145.6 (89.5) | 149.0 (74.5) | 163.8 (84.7) | 0.258 |
| Scond (1/kL) | 24.2 (20.6) | 23.8 (16.0) | 29.8 (14.2) | 28.3 (21.9) | 0.047 |

Missing data included 40 records for LCI and FRC, 42 records for Sacin and 56 records for Scond.

**Table S3.** Results from multivariable linear regression models for average Scond (1/kL) excluding Morwell as a predictor

| Predictors | Unweighted and unimputed model |  | Unweighted and imputed model |  | Weighted and imputed model |  |
| --- | --- | --- | --- | --- | --- | --- |
|  | Coef (95% CI) | p-value | Coef (95% CI) | p-value | Coef (95% CI) | p-value |
| Exposure to PM<sub>2.5</sub> | **2.15 (0.67, 3.63)** | **0.004** | **2.06 (0.62, 3.50)** | **0.005** | 1.47 (-0.12, 3.05) | 0.070 |
| Age (per 10 years) | **2.73 (1.43, 4.03)** | **<0.001** | **2.73 (1.45, 4.01)** | **<0.001** | **2.38 (1.16, 3.61)** | **<0.001** |
| Height | 0.07 (-0.21, 0.35) | 0.628 | 0.05 (-0.23, 0.32) | 0.732 | 0.01 (-0.27, 0.28) | 0.958 |
| Weight | 0.07 (-0.03, 0.17) | 0.188 | 0.07 (-0.03, 0.18) | 0.161 | 0.10 (0.02, 0.19) | 0.020 |
| Male | -2.10 (-7.41, 3.21) | 0.437 | -1.42 (-6.77, 3.93) | 0.601 | -0.43 (-5.45, 4.59) | 0.866 |
| Smoking status |  |  |  |  |  |  |
| Non-smoker | Ref |  | Ref |  | Ref |  |
| Ex-smoker | 3.11 (-0.74, 6.96) | 0.113 | 2.86 (-0.93, 6.65) | 0.138 | 3.37 (-0.60, 7.34) | 0.096 |
| Current smoker | 0.83 (-4.44, 6.09) | 0.758 | 1.43 (-3.74, 6.60) | 0.588 | 2.41 (-2.21, 7.03) | 0.305 |
| Spirometric COPD or self-reported asthma attack in the last 12 months | 3.15 (-0.35, 6.65) | 0.078 | 2.49 (-0.97, 5.94) | 0.158 | 1.70 (-1.71, 5.11) | 0.326 |
| Employed | -0.50 (-4.38, 3.38) | 0.800 | -0.55 (-4.40, 3.30) | 0.778 | -1.42 (-5.02, 2.17) | 0.437 |
| Certificate, University or other Tertiary Institute degree | 1.97 (-1.63, 5.58) | 0.283 | 1.99 (-1.47, 5.44) | 0.259 | 1.90 (-1.75, 5.56) | 0.306 |
| Occupational exposure | -2.01 (-5.91, 1.89) | 0.313 | -2.40 (-6.29, 1.48) | 0.224 | -1.03 (-4.72, 2.66) | 0.582 |

Note: all coefficients were estimated from multivariate linear regression adjusted for age, gender, height, weight, employment, education, smoking status, asthma and COPD status and occupational exposure. Missing data included 56 records for Scond and 6 records for education level.
Abbreviation: Scond = conductive ventilation heterogeneity

**Table S4.** Multivariable model of Sacin (1/kL) associated with 10 μg/m^3^ increase in mean mine fire related PM_2.5_ exposure excluding Morwell as a predictor (unweighted and unimputed)

| **Predictors** | **Coef (95% CI)** | **p-value** |
| --- | --- | --- |
| Exposure to PM<sub>2.5</sub> (per 10 µg/m<sup>3</sup>) | -2.15 (-7.83, 3.52) | 0.457 |
| Age (per 10 years) | **19.8 (14.9, 24.8)** | **<0.001** |
| Height | -0.90 (-1.96, 0.16) | 0.095 |
| Weight | 0.15 (-0.24, 0.53) | 0.451 |
| Male | **23.4 (3.26, 43.6)** | **0.023** |
| Smoking status |  |  |
| Non-smoker | Ref |  |
| Ex-smoker | **31.9 (17.2, 46.6)** | **<0.001** |
| Current smoker | **55.9 (35.8, 76.1)** | **<0.001** |
| Spirometric COPD or self-reported asthma attack in the last 12 months | **31.1 (17.7, 44.5)** | **<0.001** |
| Employed | -9.13 (-23.9, 5.66) | 0.226 |
| Certificate, University or other Tertiary Institute degree | -6.64 (-20.3, 7.04) | 0.341 |
| Occupational exposure | -1.37 (-16.1, 13.4) | 0.855 |

Note: Coefficients were estimated from unweighted and unimputed multivariate linear regression. Missing data included 42 records for Sacin and 6 records for education level.
Abbreviation: Sacin = acinar ventilation heterogeneity

## Notes

### Competing Interest Statement

The authors have declared no competing interest.

### Funding Statement

This study was funded by the Department of Health, State of Victoria, Australia.

### Author Declarations

Monash University Human Research Ethics Committee of Monash University gave ethical approval for this work (project number: 1078) The Alfred Ethics Committee of the Alfred Hospital gave ethical approval for this work (project number: 90/21).

